# Extended laboratory panel testing in the Emergency Department for risk-stratification of patients with COVID-19: a single centre retrospective service evaluation

**DOI:** 10.1101/2020.10.06.20205369

**Authors:** Mark J Ponsford, Ross J Burton, Leitchan Smith, Palwasha Khan, Robert Andrews, Simone Cuff, Laura Tan, Matthias Eberl, Ian R Humphreys, Farbod Babolhavaeji, Andreas Artemiou, Manish Pandey, Stephen Jolles, Jonathan Underwood

## Abstract

**Background:** The role of specific blood tests to predict poor prognosis in patients admitted with infection from SARS-CoV2 virus remains uncertain. During the first wave of the global pandemic, an extended laboratory testing panel was integrated into the local pathway to guide triage and healthcare resource utilisation for emergency admissions. We conducted a retrospective service evaluation to determine the utility of extended tests (D-dimer, ferritin, high-sensitivity troponin I, lactate dehydrogenase, procalcitonin) compared to the core panel (full blood count, urea & electrolytes, liver function tests, C-reactive protein).

**Methods:** Clinical outcomes for adult patients with laboratory-confirmed COVID-19 admitted between 17^th^ March to 30^st^ June 2020 were extracted, alongside costs estimates for individual tests. Prognostic performance was assessed using multivariable logistic regression analysis with 28-day mortality used as the primary endpoint, and a composite of 28-day intensive care escalation or mortality for secondary analysis.

**Results:** From 13,500 emergency attendances we identified 391 unique adults admitted with COVID-19. Of these, 113 died (29%) and 151 (39%) reached the composite endpoint. “Core” test variables adjusted for age, gender and index of deprivation had a prognostic AUC of 0.79 (95% Confidence Interval, CI: 0.67 to 0.91) for mortality and 0.70 (95% CI: 0.56 to 0.84) for the composite endpoint. Addition of “extended” test components did not improve upon this.

**Conclusion:** Our findings suggest use of the extended laboratory testing panel to risk stratify community-acquired COVID-19-positive patients on admission adds limited prognostic value. We suggest laboratory requesting should be targeted to patients with specific clinical indications.

## Introduction

In December 2019, a novel coronavirus disease (COVID-19) was reported in China, caused by severe acute respiratory syndrome coronavirus 2 (SARS-CoV-2) (1). In the first 8 months since its emergence, SARS-CoV-2 has caused over 32 million infections and more than a million deaths worldwide (2). The majority of patients with COVID-19 experience a mild influenza-like illness, however approximately 15-25% of those admitted to hospital develop pneumonia that may evolve into acute respiratory distress syndrome (ARDS) (3-6). Experience from the Italian region of Lombardy highlighted the potential of uncontrolled COVID-19 outbreaks to rapidly overwhelm local intensive care capacity and healthcare systems (5). In the United Kingdom, Wales has one of the lowest number of intensive care beds per head of population in Europe (7, 8), prompting implementation of scoring systems to support patient triage and allocation of healthcare resources.

The ability to identify patients at greatest risk of developing life-threatening complications from SARS-CoV-2 infection based on haematological and biochemical laboratory markers was suggested early in the pandemic. A range of admission tests including D-dimer, ferritin, high-sensitivity troponin I (hs-Trop), and lactate dehydrogenase (LDH) have been linked with disease severity and risk of death (9-13). Similar findings have been replicated in meta-analysis (14). Furthermore, use of a broader range of laboratory tests in patients with COVID-19 has been supported by the UK Royal College of Pathologists (15). Accordingly, an extended panel of laboratory tests was integrated within the standard of care pathway for COVID-19 admissions presenting via the Emergency Department (ED) of the University Hospital of Wales. This panel consisted of both “core” (full blood count, FBC; urea & electrolytes, U&E; liver function tests, LFTs; C-reactive protein, CRP) and “extended” test components (D-dimer; LDH; ferritin; hs-Trop; and procalcitonin, PCT).

A joint National Health Service (NHS)-University collaboration supporting the rapid creation of an electronic healthcare registry (see extended methods) provided a timely opportunity to retrospectively assess the value and cost of implementing this extended laboratory panel. This is particularly relevant given a recent systematic review of methodological and reporting standards highlighting caution before extrapolating models and decision thresholds derived from prognostic biomarker studies into local clinical practice (18). The role of extended components remains poorly represented in prognostic studies within the UK population to date (4, 16-19). We therefore conducted a service evaluation focusing on the ability of these tests to predict mortality or escalation to intensive care in the first 28-days following admission, in adult patients with PCR-confirmed COVID-19. Our primary aim was to assess how addition of components of the extended panel altered the prognostic performance of the core panel (15). Our secondary aim was to explore the additional cost of extended testing components. Together, this directs refinement of a risk stratification panel with potential cost savings ahead of future waves of COVID-19.

## Methods

### Study population

We identified patients aged ≥ 18 years admitted between 17^th^ March 2020 to 30^th^ June 2020 via the Emergency Department (ED) of the University Hospital of Wales (Cardiff, UK). This 1,000-bed hospital is a tertiary referral centre within the region with the greatest recorded total of COVID-19 case positives in Wales (20). Only patients with SARS-CoV-2 infection confirmed by positive reverse transcriptase polymerase chain reaction (PCR) on nasopharyngeal swab, and likely community-acquired disease (defined as swab positive between 14 days prior or 7 days following the date of initial emergency attendance) were included. Patients transferred in from other hospitals were excluded.

The primary dataset was extracted as part of a service evaluation to assist local care planning. A fully anonymised dataset was created by a member of the Health Board NHS IT team. Prior to anonymisation, the postcode was used to extract the Welsh Index of Multiple Deprivation (WIMD) for each patient, as obtained from https://wimd.gov.wales/. As such, ethical approval was not required for this study.

Data fields including admission date, clinical outcomes, and laboratory measurements were integrated into an electronic healthcare registry “Cardiff Hospital Admissions Database” (CHAD) using a bespoke software package: CHADBuilder (*see extended methods*). Laboratory test results from the index presentation reported within the first 72 hours of ED presentation were considered as candidate variables.

### Outcomes

28-day mortality was chosen as the primary endpoint in accordance with UK COVID-19 mortality reporting. To support generalisability between studies (4, 16) we performed secondary analysis using the composite endpoint of 28-day mortality or admission to intensive care.

### Laboratory testing panel

All testing was performed in the United Kingdom Accreditation Service (UKAS)-accredited Biochemistry, Immunology, and Haematology Laboratories at the University Hospital of Wales. Cost estimates were obtained from the Health Board Laboratory Medicine Directorate, reflecting consumables, reagent, analyser running and maintenance costs, and staff time chargeable to NHS test requestors.

### Statistical analysis

Statistical significance testing was performed according to the data encountered: for categorical data, such as gender, Fisher’s exact or chi-square testing was performed. For continuous data, Welch’s t-tests were used if the assumptions of normality were met; otherwise non-parametric Mann-Whitney U tests were employed. In edge-cases, permutation testing was performed. Two-sided statistical significance was set at p <0.05, with Bonferroni correction for multiple testing.

### Model development

Candidate laboratory variables were triaged for inclusion based on their membership of core or extended laboratory test panels, before a data-driven approach was applied. This included assessment of variability, individual p-values corrected for multiple comparisons and multi-collinearity with generation of a Spearman’s rank correlation matrix.

Logistic regression, support vector machines, random forest, and gradient boosted trees were all considered for multivariate predictive models. Models with complexity greater than logistic regression were found to offer little improvement (data not shown). Multivariate logistic regression was implemented in Python (version 3.7) using the Scikit-Learn package (version 0.23) (21) and Statsmodels (version 0.11). Complete case analysis was conducted to enable meaningful comparison between core and extended tests.

To minimise bias, models were evaluated using cross-validation with 5-folds, with stratification to account for class imbalance. Performance statistics are reported as the average across all folds with binomial proportion 95% confidence intervals. Model discrimination was assessed by area under receiver operating characteristic curve (ROC AUC), accuracy (balanced by support) and weighted F1 score (the average F1 score was calculated for each class and weighted by support). In addition to these performance metrics, threshold-performance curves were generated to assess the effect of the decision threshold on model sensitivity and specificity (22). Source code for all models can be found on GitHub: https://github.com/burtonrj/CardiffCovidBiomarkers

Our evaluation is reported using the transparent reporting of a multivariable prediction model for individual prognosis or diagnosis (TRIPOD) guidance for Prediction Model development and validation (See Appendix).

### Patient involvement

These data were generated as part of a rapid service improvement and as such patients were not involved in the setting of the research question or interpretation of the study.

## Results

### Definition and overview of service evaluation cohort

We focused on admissions occurring after the operational roll-out of the first extended laboratory test panel components into standard clinical practice. During this 105 day period, over 13,500 ED attendances were recorded. Of these, 391 adults were admitted via ED with a laboratory-confirmed diagnosis of COVID-19 meeting our definition of likely community-acquired COVID-19 (**Figure 1: Study Flowchart**). The median age was 69 years (interquartile range, IQR: 55 - 75 years) with males predominant (52.4%). Within 28-days of index ED attendance, 113 deaths occurred (29% mortality), and 151 patients reached the composite secondary endpoint of intensive care admission/death (39%).

**Figure 1:**
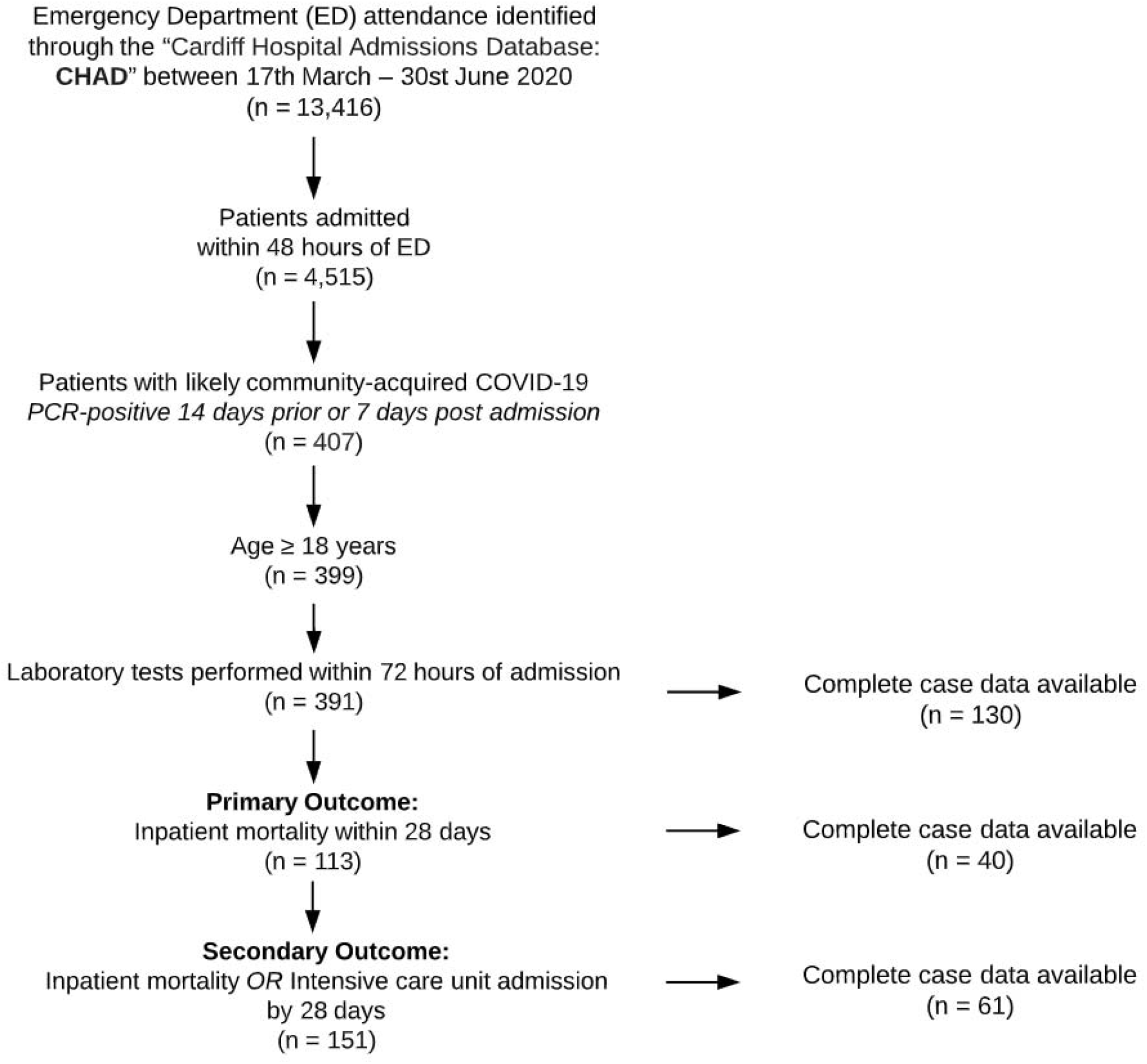
Study Flowchart.

### Univariate analysis of laboratory predictors of adverse inpatient course

We next analysed the association between individual candidate variables and patient outcomes to identify important predictors of adverse outcome. Admission clinical variables are presented in **Table 1** (**for full dataset, Supplementary S1&S2)**. Advanced age was strongly associated with increased risk of death and the composite of ICU admission and death. In contrast, neither gender nor socio-economic deprivation were associated with 28-day mortality. For laboratory variables, missing data were rare for core test panel components. Within the extended testing panel, hs-Trop and D-dimer were available in 70-80% of patients admitted with COVID-19 within the first 72 hours of ED attendance. An early admission PCT test result was available in 40% of patients, whilst ferritin and LDH levels were recorded in 42-46% of cases. Testing rates were similar between patient survival groups.

**Table 1:** Demographic and selected clinical laboratory predictor variables on admission for evaluation cohort. Variables captured in the summarised cohort of community-acquired PCR-confirmed COVID-19 cases admitted through the ED between 17^th^ March 2020 and 30^th^ June 2020. Summary statistics are given as the median and range for continuous variables and absolute counts for discrete variables. * Welsh index of multiple deprivation, WIMD, is ranked from 1 (most deprived) to 1,909 (least deprived), and presented as frequencies within each quartile. ^†^Fischer’s exact test.

|  | Survivors (n= 278) |  | Non-survivors (n=113) |  |  |
| --- | --- | --- | --- | --- | --- |
| Variable | Frequency | % Missing | Frequency | % Missing | p-value |
| **Gender** |  | 0 |  | 0 |  |
| Male | 141 (50.7%) | - | 64 (56.6%) | - | 0.316† |
| Female | 137 (49.3%) | - | 49 (44.4%) | - |  |
| **WIMD\*** |  | 2.52 |  | 0.885 | 0.228† |
| Quartile 1 (< 246) | 65 (23.4%) | - | 29 (25.6%) | - |  |
| Quartile 2 (246 – 871) | 61 (21.9%) | - | 35 (31.0%) | - |  |
| Quartile 3 (872 – 1672) | 73 (26.3%) | - | 23 (20.4%) | - |  |
| Quartile 4 (> 1672) | 72 (25.9%) | - | 25 (22.1%) | - |  |
|  | **Median [IQR]** |  | **Median [IQR]** |  |  |
| Age, years | 63.5 [51.25 - 77.75] | 0 | 81.0 [71.0 - 88.0] | 0 | < 0.0001 |
| **“Core” test component** |  |  | Median [IQR] |  |  |
| Albumin, g/L | 33.0 [29.0 - 36.0] | 3.24 | 29.0 [26.0 - 32.0] | 1.77 | < 0.0001 |
| Alkaline phosphatase U/L | 80.0 [63.0 - 111.5] | 0.04 | 100.0 [76.0 - 133.5] | 0.02 | 0.0006 |
| Alanine transaminase U/L | 27.0 [17.0 - 46.0] | 0.04 | 23.0 [14.0 - 32.0] | 0.02 | 0.256 |
| Bilirubin µmol/L | 10.0 [7.0 - 15.0] | 0.04 | 12.0 [8.0 - 17.0] | 0.02 | 0.0018 |
| C-reactive protein, mg/L | 70.5 [21.0 - 131.75] | 0.00 | 98.0 [55.75 - 164.75] | 2.65 | 0.005 |
| Creatinine, µmol/L | 82.0 [66.0 - 105.0] | 0.36 | 111.0 [79.0 - 192.0] | 0.00 | < 0.0001 |
| Estimated GFR ml/min/1.73m<sup>2</sup> | 75.0 [55.0 - 89.0] | 0.01 | 51.0 [25.0 - 74.0] | 0.00 | < 0.0001 |
| Globulin g/L | 38.0 [34.0 - 42.0] | 0.07 | 40.0 [36.0 - 45.75] | 0.06 | 0.0161 |
| Haemoglobin g/L | 135.0 [122.0 - 149.0] | 0.00 | 135.0 [122.0 - 149.0] | 0.00 | 0.036 |
| Lymphocyte count x10<sup>9</sup>/L | 1.0 [0.7 - 1.4] | 0.00 | 0.9 [0.6 - 1.22] | 0.01 | 0.961 |
| Neutrophil count x10<sup>9</sup>/L | 5.4 [3.7 - 7.98] | 0.00 | 7.3 [4.57 - 9.8] | 0.01 | 0.017 |
| Neutrophil : Lymphocyte ratio | 5.25 [3.25 - 9.81] | 0.36 | 8.11 [4.34 - 14.53] | 0.88 | 0.011 |
| Platelet count x10<sup>9</sup>/L | 234.0 [183.75 - 294.5] | 0.00 | 216.0 [160.0 - 285.0] | 0.00 | 0.210 |
| Potassium mmol/L | 3.9 [3.6 - 4.3] | 0.01 | 4.1 [3.73 - 4.6] | 0.03 | 0.0006 |
| Protein g/L | 71.0 [67.0 - 76.0] | 0.07 | 70.0 [65.0 - 74.0] | 0.06 | 0.04 |
| Sodium mmol/L | 137.0 [134.0 - 139.0] | 0.00 | 138.0 [134.0 - 141.0] | 0.00 | < 0.0001 |
| Urea mmol/L | 5.7 [4.0 - 8.5] | 0.36 | 10.2 [7.2 - 16.2] | 0.00 | < 0.0001 |
| White blood cell count x10<sup>9</sup>/L | 7.35 [5.3 - 9.93] | 0.00 | 9.0 [6.4 - 12.3] | 0.00 | 0.016 |
| **“Extended” test component** |  |  |  |  |  |
| Ferritin µg/L | 325.0 [125.0 - 828.0] | 57.9 | 482.0 [245.5 - 993.5] | 54.9 | 0.395 |
| High sensitivity D-dimer µg/L | 926.5 [587.75 - 1750.0] | 28.1 | 1497.0 [929.0 - 3885.0] | 30.1 | 0.0003 |
| High Sensitivity Troponin I ng/L | 7.0 [3.0 - 23.0] | 24.8 | 35.5 [15.75 - 117.0] | 22.1 | < 0.0001 |
| Lactate dehydrogenase U/L | 349.5 [270.25 - 549.75] | 53.2 | 383.5 [290.5 - 501.25] | 54.0 | 0.999 |
| Procalcitonin µg/L | 0.14 [0.06 - 0.38] | 41.4 | 0.31 [0.09 - 0.86] | 38.9 | 0.019 |

In univariate analysis of the core laboratory panel components, increased CRP, neutrophil:lymphocyte ratio, urea, creatinine; or decrease in serum albumin were all strongly associated with risk of death. Within the extended panel, D-dimer, hs-Trop and PCT differed between survivors and non-survivors on univariate analysis (**Figure 2; Supplementary S3**). No extended panel members were associated with development of the composite outcome (**Supplementary S4&S5**). Age was associated with several variables (**Supplementary S6**), indicating it could confound the relationship between a test result and mortality.

**Figure 2:**
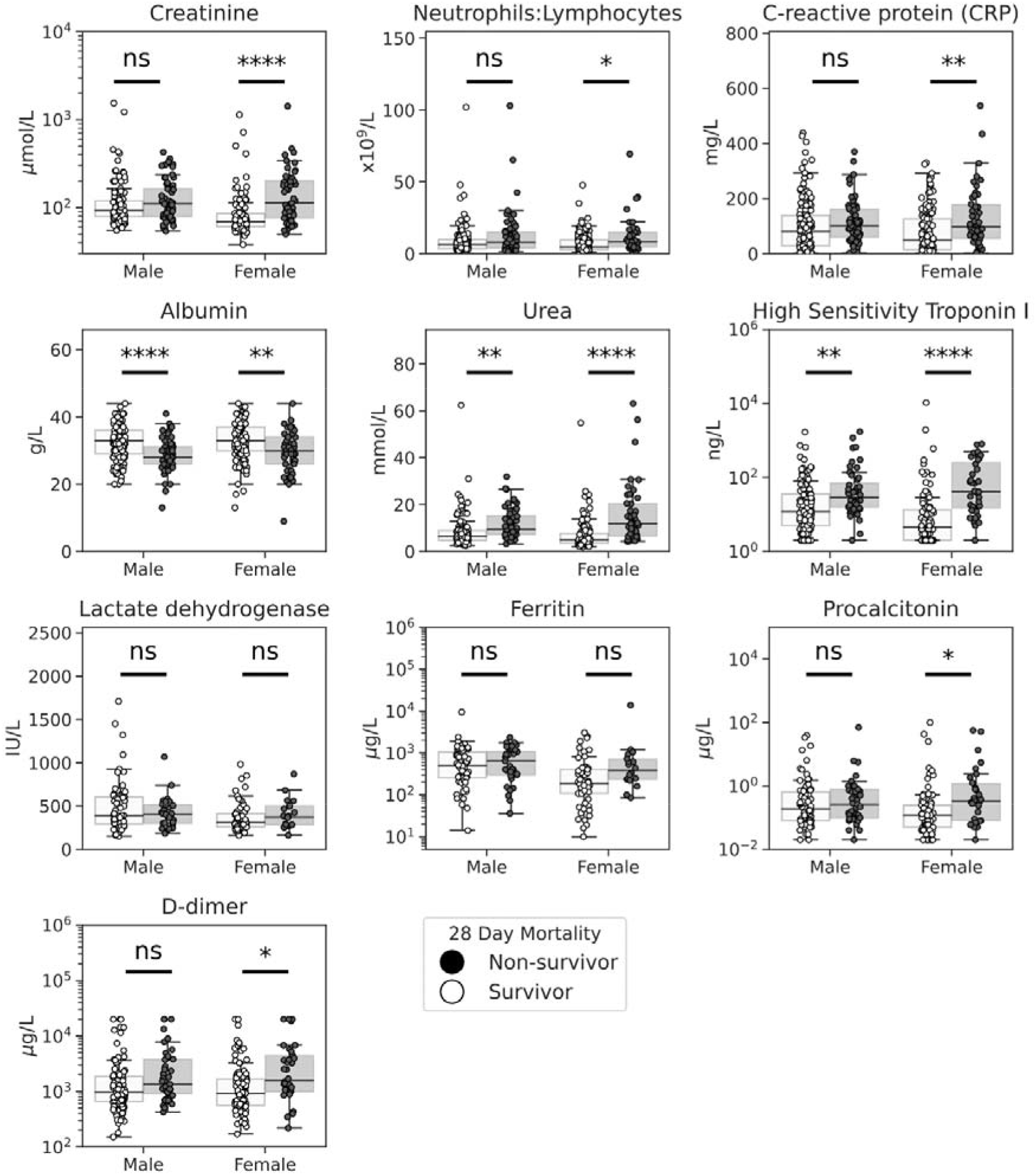
Laboratory test results according to survival outcome and grouped by gender. Box and swarm plots showing the initial laboratory test results from laboratory-confirmed COVID-19 patients, grouped by gender and 28-day mortality. Example variables considered from the components of the core laboratory test panel. * indicates level of significance, assessed by Mann-Whitney U test with correction for multiple testing: * p<0.05, ** p<0.01, *** p<0.005. **** p <0.001.

### Development of prognostic model based on core and extended laboratory admission test panels

We therefore used multivariate logistic regression to adjust for the role of age, whilst controlling for gender and WIMD, based on consistent identification of their contribution to outcomes in COVID-19 cohorts (23, 24). Restricting to cases with complete data (n=130) across core and extended laboratory tests, we found an optimal combination of core test variables to be CRP, albumin, urea, neutrophil:lymphocyte ratio, creatinine, age, gender and WIMD (**Figure 3, Supplementary S7**). This gave a prognostic AUC of 0.79 (95% confidence interval, CI: 0.67 to 0.91) for 28-day mortality, and 0.70 (95% CI: 0.56 – 0.84) for the composite outcome.

**Figure 3:**
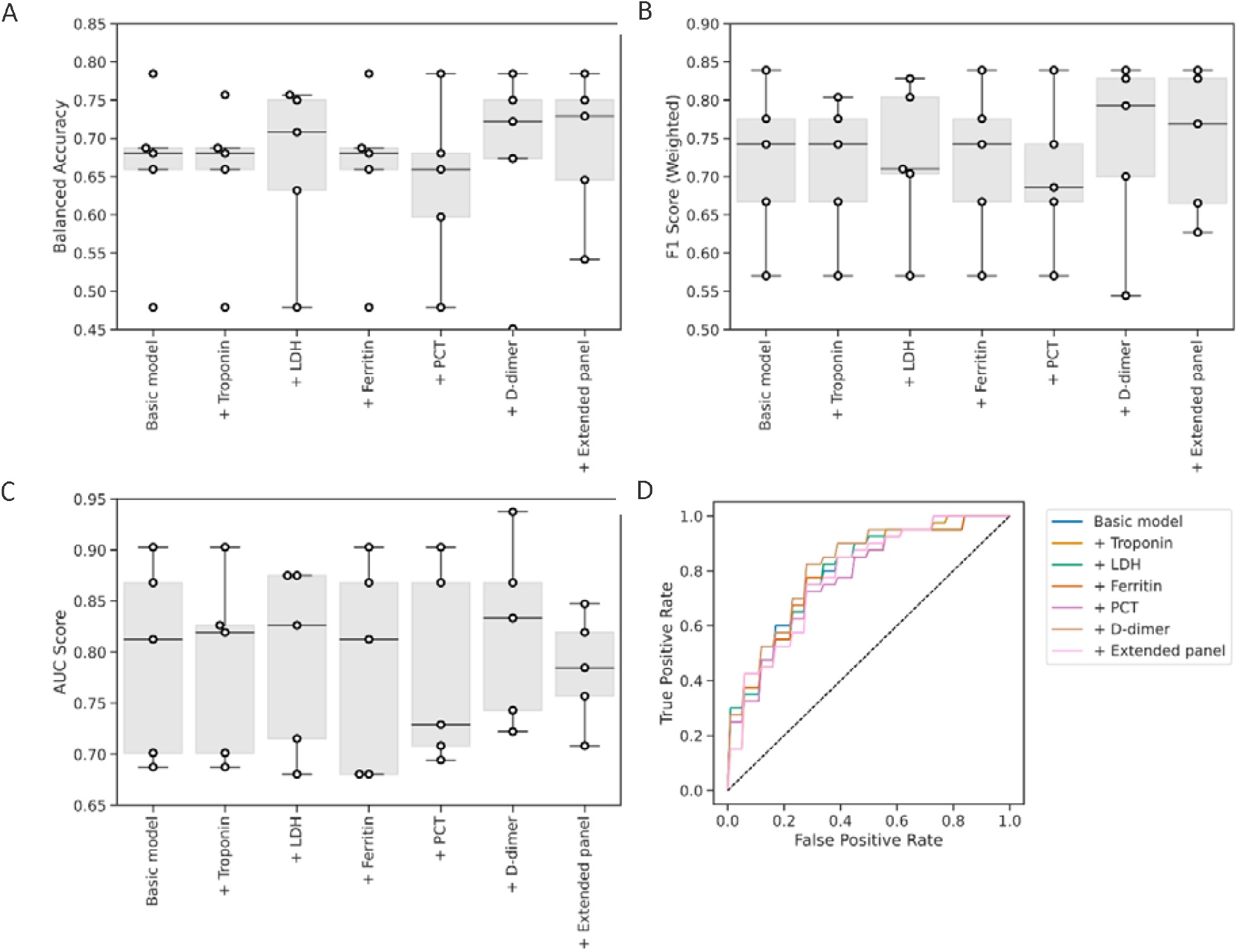
Balanced accuracy (A), weighted F1 score (B), AUC score (C) and ROC curves (D) for models with sequential inclusion of extended biomarkers for prediction of 28 day mortality.

We next assessed the discriminative value associated with inclusion of extended panel components within our multivariate model, relative to this core test set (**Figure 5**). Addition of D-dimer resulted in a marginal increase in mean AUC score to 0.82, but this was not significantly different (95% CI: 0.71 – 0.85) to the performance of core testing alone. Concerning the composite outcome, addition of admission hs-Trop to the core panel resulted in the greatest AUC score: 0.72 (95% CI: 0.60 to 0.83) but again, this did not represent a significant increase in performance to the core panel alone (**Supplementary S7&S8)**. Consideration of extended test components individually or in combination did not improve upon this. To internally validate these findings, we performed stratified cross-validation, observing convergence of training and validation curves, thus suggesting a low-risk of over-fitting associated with these models (**Supplementary S9&S10**). Assessing the calibration of these models across a range of performance metrics by varying the decision threshold (the probability at which a patient is predicted to either die or be admitted to intensive care), we found no significant benefit from addition of the extended relative to the core laboratory testing panel (**Supplementary S11&12**).

### Patterns of extended panel requesting during the first wave

Local cost estimates for NHS requesting the core laboratory panel totalled £16.44 per patient, with an additional £55.48 incurred for the extended set (**Supplementary S13**). In order to contextualise testing beyond the cohort of community-acquired COVID-19, we constructed a run-chart of test requesting within the first 72 hours of admission via ED and COVID-19-related admissions (**Figure 4)**. D-dimer and hs-Trop testing rates rose in line with COVID-19 admissions during March and April, with a 1-2 week delay apparent for LDH, ferritin, and PCT requesting. Strikingly, whilst COVID-19 admissions declined following the April peak, the intensity of extended biomarker panel requesting remained. Using January and June 2020 to represent requesting patterns before and after the first wave of COVID-19, mean monthly requesting increased by 29.7%, 224%, and 588% for hs-Trop, ferritin, and LDH, respectively. In contrast, recorded monthly ED attendance fell by 24.0% over this period. PCT and quantitative D-dimer were specifically introduced in response to the pandemic, but still averaged >50 daily test requests within the early admission period during June. Across the evaluation period, over 6,400 D-dimer and 5,400 PCT requests were made, with an estimated service cost of £246,000.

**Figure 4:**
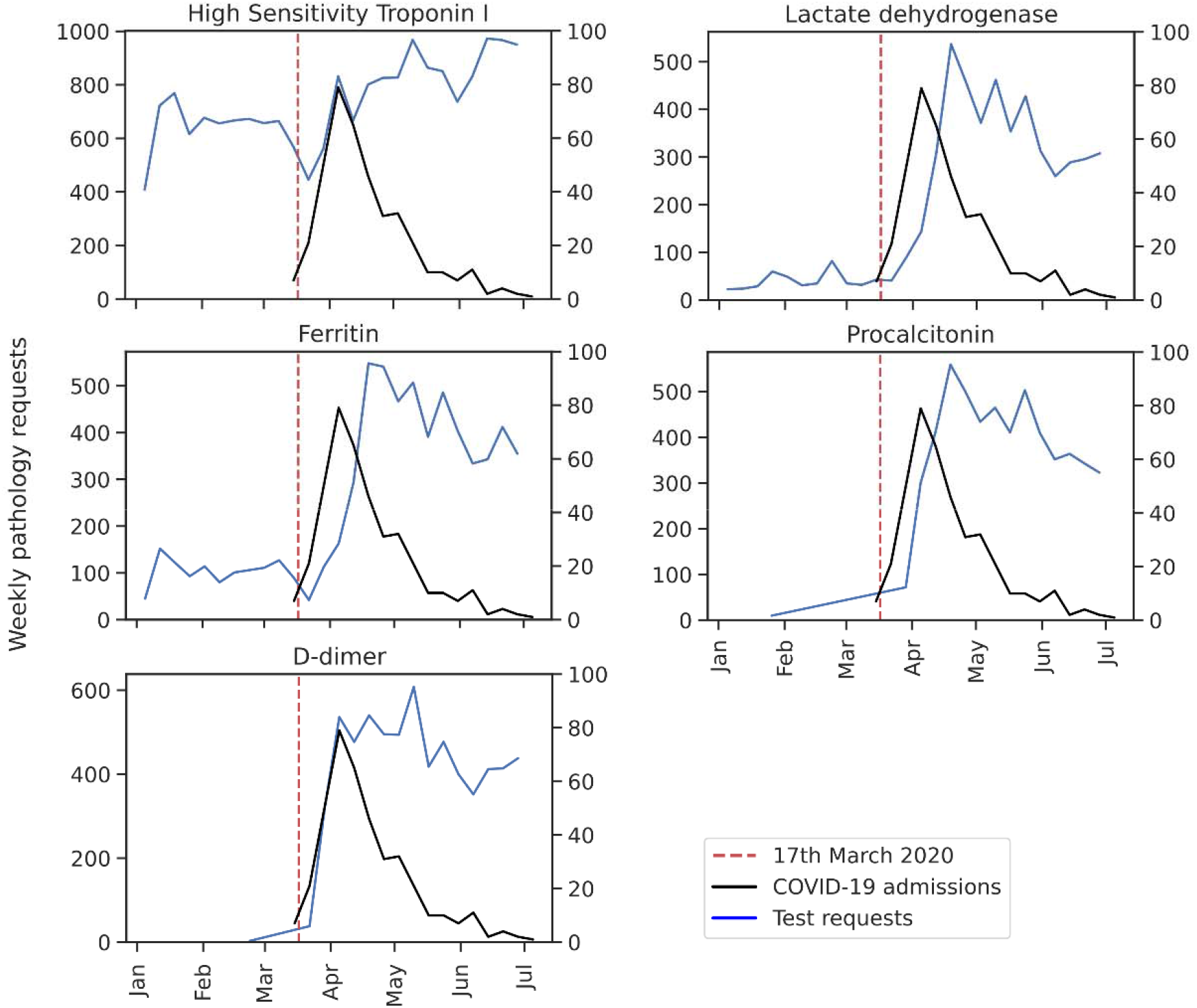
Daily COVID-19 admission rates and test requesting patterns during the early admission period. Run-chart showing Emergency Unit (ED) admission rates for patients with confirmed laboratory-confirmed COVID-19 (right y-axis, black), and accompanying tests performed within 72 hours of ED attendance (left y-axis, blue) during the first wave of the COVID-19 pandemic. The dotted line indicates the roll-out of extended panel testing from 17^th^ March 2020.

## Discussion

To support the effective and efficient use of resources through evidence-based clinical practice, we conducted a service evaluation determining the prognostic value associated with routinely performed laboratory investigation in 130 adults admitted with community acquired SARS-CoV-2 infection. By leveraging a bespoke electronic healthcare registry, we reveal an extended panel (including D-dimer, LDH, hs-Trop, ferritin, PCT) provided only limited additional prognostic information beyond that provided by components of the core panel (FBC, U&E, LFT, and CRP). Together, this directs refinement of the clinical testing panel employed before and during future potential waves, underlining the relevance of this registry-approach to support cost-utility of investigation pathways.

We identified 5 studies within the peer-reviewed and pre-print literature concerning laboratory biomarker risk stratification of adult COVID-19 admissions in the UK population (4, 16-19). The largest reported, an 8-point pragmatic risk score developed by the ISARIC Consortium, achieved a modest AUC performance score of 0.77 (95% CI 0.76 to 0.77) when predicting 28-day mortality (18). To date, only 1 UK study has considered the prognostic role of variables within our extended laboratory panel (16). In their prospective analysis of 155 patients, Arnold et al. found conventional laboratory biomarkers such as CRP and neutrophil elevation offered limited prognostic performance (with AUC scores of 0.52 and 0.54, respectively), whilst ferritin, PCT, hs-Trop, and LDH performed with AUC scores of 0.65 to 0.71. It is important to note that within this study cohort, the incidence of clinical deterioration was low (overall mortality was only 4% vs 29% for our service evaluation) which may have limited the power of the study (16). This highlights regional variation in rates of hospitalisation and mortality, and further motivated a locally-led assessment of practice.

Consistent with the emerging COVID-19 literature, we observed an association between laboratory markers of acute phase inflammatory response (elevated neutrophil count, CRP; depressed lymphocytes and albumin), cardiac injury, activation of thrombosis, and renal impairment with subsequent adverse outcome (6, 23, 25). We found a combination of CRP, albumin, urea, neutrophil:lymphocyte ratio, and creatinine alongside simple demographics achieved an AUC of 0.79 (95% CI: 0.67 – 0.91) when predicting 28-day mortality, and 0.70 (95% CI: 0.56 – 0.84) for the composite endpoint. We found no evidence that use of this panel at admission significantly improved performance for either outcome. Importantly, we identified use of the extended laboratory panel continued despite falling rates of COVID-19 presentations, indicating a change in routine test requesting patterns. Addition of the extended laboratory test panel equates to £54 per patient (a relative cost increase of over 400% to the core panel alone), with significant cost ramifications when performed at scale.

Our service evaluation has several strengths, notably assessment of the performance of an extended panel of laboratory tests not widely considered in UK prognostic studies to date (4, 16-19). These tests were integrated into routine practice prior to the local peak of the pandemic, based on available literature and national guidelines (15). In contrast with the batched analysis undertaken under research condition in previous studies (16), all tests described here were conducted by accredited laboratories using platforms calibrated to international reference standards, facilitating future data sharing. Our multivariate approach is well-suited to investigate whether specific laboratory tests provide additional prognostic value beyond conventional parameters (26), using inclusion criteria and clinically-relevant endpoints in line with other reported studies (4, 17, 18). Finally, we considered the service costs that accompanied implementing these tests into routine practice (27), a relevant factor often neglected in other publications.

Our evaluation also has a number of limitations, reflecting the challenges of clinical data collection during an epidemic. It represents retrospective experience from a single tertiary referral centre, limiting sample size and the generalisability of our findings. Secondly, availability of extended test panel results during the early admission period was mixed. Admission D-dimer and hs-Trop results were available for 70-80% of patients, comparing favourably to a similar UK registry-based study where D-dimer results were only available at time of admission in 37.2% (17). Conversely, we observed high rates of missing data for LDH, ferritin, and PCT, undermining their relevance as a prognostic tool. This was likely due to operational factors such as a delay in test roll out relative to epidemic peak, and requirement for an additional sample tube. Because it cannot be assumed that data are missing at random, we chose to perform complete case analysis. Although this limits our statistical power, it avoids unfounded assumptions and potentially invalid imputation. In its current form, the CHAD-registry lacks detailed information on patient-level physiological observations, nature of co-morbidities, and therapeutic interventions. Similarly, all registry-linked laboratory values were available to clinicians, and are likely to have influenced management decisions. With advances in clinical care diagnostics, therapeutics are likely to alter the observed performance of the prognostic model. These limitations apply equally to pragmatic risk-scores (4, 16, 18). Finally, we recognise our evaluation consider the index test result and a specific question of inpatient prognosis, and additional indications exist for requesting components of the extended laboratory tests that fall outside of our primary and secondary endpoints. For instance, the use of PCT is often employed to support antibiotic stewardship (28), and was integrated into routine practice locally in early April 2020. There may also be merit in more targeted use of additional testing particularly as therapeutic options evolve. Hence, whilst we highlight the significant associated healthcare costs with implementation of extended laboratory testing, we do not make specific claims concerning the potential savings from discontinuing unnecessary investigations (27).

### Implications for practice

Laboratory markers supporting early risk stratification of patients are often used in the ED setting, and have been shown to benefit patient triage (29). Our data suggest that systematic testing of COVID-19-positive patients upon admission with an “extended” laboratory panel provide little additional prognostic information for COVID-19 mortality or intensive care admission “core” tests. Besides the financial impact, over-requesting of laboratory tests are likely to increase the number of false-positive results, with the potential to lead to further potentially harmful tests (e.g. computed tomography pulmonary angiography in patients with marginally elevated D-dimer but no clinical indication of thromboembolic disease) (30). We suggest that the use of these laboratory markers be targeted to patients with specific clinical indications for these, such as PCT to guide antibiotic prescription or hs-Trop in patients with suspected myocardial injury.

In conclusion, we report our real-world experience from the use of an extended laboratory prognostic testing panel in patients hospitalised with community-acquired COVID-19. These findings directly inform clinical practice, guiding cost-efficient use of resources in potential future waves.

## Data Availability

Requests for data sharing will be reviewed by a clinical and information regulatory governance panel and considered on an individual basis.

https://github.com/burtonrj/CardiffCovidBiomarkers

## Author contributions

MJP and SJ conceived the service evaluation. MJP wrote the first draft, supervised by SJ, and JU. LS, RA and RJB performed initial data extraction. RJB constructed CHAD, and led data analysis support for multivariate modelling from AA and PK. All authors contributed scientific and clinical guidance regarding design, feature selection, and have reviewed the final draft.

## Acknowledgements

We thank James Webb and Philip Clee (NHS IT governance) for their advice and guidance in facilitating the Electronic Healthcare Registry underpinning this service evaluation, and Nigel Roberts and Alun Roderick for assistance in providing costings for laboratory tests. Additional support was provided by Professor Valentina Escott-Price, Dr Martyn Stones and Dr Irina Erchova.

## Key Messages

During the first wave of the pandemic, the literature and guidance from the UK Royal College of Pathologists supported the use of extended biochemistry and haematology testing upon admission to support risk stratification of patients with COVID-19 infection-however the prognostic performance of these markers remains unclear.

Our service evaluation suggests that systematic testing of COVID-19-positive patients with likely community-acquired disease upon admission with an “extended” laboratory panel (high-sensitivity Troponin I, Ferritin, Lactate Dehydrogenase, procalcitonin, or quantitative D-dimer) provides limited additional prognostic information for 28-day mortality or intensive care admission, relative to conventional “core” tests such as a full blood count, renal function, C-reactive protein combined with simple demographics.

Few clinicians know the cost of the tests they request for their patients. With individual “extended” panel members costing over £20 per test, and thousands of tests requested per month within a single hospital, these costs quickly escalate.

Besides the financial impact, over-requesting of laboratory tests are likely to increase the number of false-positive results, with the potential to lead to further potentially harmful investigations (e.g. computed tomography pulmonary angiography in patients with marginally elevated D-dimer but no clinical indication of thromboembolic disease). We suggest that the use of these laboratory markers be targeted to patients with specific clinical indications for these, such as procalcitonin to guide antibiotic stewardship or hs-Troponin I in patients with suspected myocardial injury.

