## Supplementary for "Extended laboratory panel testing in the Emergency Department for risk-stratification of patients with COVID-19: a single centre retrospective service evaluation" (from another version of this preprint: version 1, 2020-10-11)

### Supplementary methods: developing an ETL pipeline in Python to generate CHAD

In early April 2020 a data processing agreement was established between Cardiff and the Vale University Health Board (C&VHB) Information Governance Department and Cardiff University to extract and analyse data detailing patient hospital events (e.g. admissions and discharge), radiology, blood tests, and demographics. This data was pseudo-anonymised on NHS servers by NHS employees and made available through a secure domain managed by NHS Wales Informatics Service (NWIS).

Data scientists at Cardiff University designed CHADBuilder, a bespoke ETL application with modules designed specifically for access to the NWIS secure domain. Extracts stored as csv files are located and retrieved from the NWIS secure domain and reconstructed on a local device or server. CHADBuilder accesses this domain by the means of a RESTful API with token authentication. Authorisation is provided through NWIS on a per-project basis. The files are checked for errors or illegal characters and then consolidated such that similar data is contained within single csv files. Once the data is validated and consolidated, an SQL database is generated on the local disk. Given the limited resources at hand, constraints that information governance poses on database hosting, and the need for a rapid solution, we chose SQLite as a database engine. This requires that some data types such as date and time be handled natively in the chosen scripting language accessing the database (e.g. Python or R) but does not require hosting and is technically easy to manage, with a single file that can be password encrypted and shared between collaborators. To help with data wrangling, dates and times are combined into an ISO 8601 standard “YYYY-MM-DD hh:mm:ss” such that they can be handled with ease in any chosen scripting language.

CHADBuilder was developed in Python version 3.7 on an Ubuntu OS version 18.04. The application itself has very few dependencies (Pandas v1.0.5, requests v2.24, tqdm v4.47, dateparser v0.7.6) and can be downloaded from our GitHub repository (https://github.com/burtonrj/CHAD). Instructions for use are simple and provided in the “readme” file. Commands result in a single SQLite database file saved to disk. The database schema diagram and a table of available variables and data types are available in the GitHub repository.

### **Supplementary S1: Clinical Laboratory data on admission for evaluation cohort for the primary endpoint (28-day mortality)**

|  | **Survivors** | | | **Non-survivors** | | |  |  |
| --- | --- | --- | --- | --- | --- | --- | --- | --- |
|  | Median [IQR] | Missing (%) | Normalised Log Variance | Median [IQR] | Missing (%) | Normalised Log Variance | **P-value** | Common language effect size |
| APTT | 31.5 [29.4 - 33.98] | 0.14 | -8.50 | 30.8 [28.2 - 34.48] | 0.10 | -8.33 | 0.538 | 0.47 |
| Alanine transaminase | 27.0 [17.0 - 46.0] | 0.04 | -6.89 | 23.0 [14.0 - 32.0] | 0.02 | -8.17 | 0.256 | 0.40 |
| Albumin | 33.0 [29.0 - 36.0] | 0.04 | -8.26 | 29.0 [26.0 - 32.0] | 0.02 | -8.78 | **< 0.0001** | 0.31 |
| Alkaline phosphatase | 80.0 [63.0 - 111.5] | 0.04 | -6.24 | 100.0 [76.0 - 133.5] | 0.02 | -5.06 | **0.0006** | 0.61 |
| Basophil count | 0.0 [0.0 - 0.0] | 0.00 | -22.42 | 0.0 [0.0 - 0.0] | 0.01 | -23.04 | 0.37 | 0.49 |
| Bilirubin | 10.0 [7.0 - 15.0] | 0.04 | -10.58 | 12.0 [8.0 - 17.0] | 0.02 | -10.19 | **0.0018** | 0.59 |
| C-reactive protein (CRP) | 71.0 [21.0 - 132.0] | 0.00 | -5.40 | 98.5 [55.75 - 168.75] | 0.03 | -5.23 | **0.0019** | 0.62 |
| Clauss fibrinogen level | 5.3 [4.1 - 6.7] | 0.14 | -11.84 | 5.0 [3.8 - 6.45] | 0.10 | -11.81 | 0.409 | 0.47 |
| Creatinine | 82.0 [65.0 - 108.0] | 0.00 | -4.32 | 111.0 [79.0 - 186.0] | 0.00 | -5.47 | **0.0082** | 0.68 |
| D-dimer | 926.5 [587.75 - 1776.0] | 0.28 | -2.88 | 1497.0 [929.0 - 3885.0] | 0.30 | -3.11 | **0.0031** | 0.66 |
| Eosinophil count | 0.0 [0.0 - 0.1] | 0.00 | -18.80 | 0.0 [0.0 - 0.1] | 0.01 | -21.24 | 0.051 | 0.45 |
| Estimated GFR | 75.0 [55.0 - 89.0] | 0.01 | -6.43 | 51.0 [25.0 - 74.0] | 0.00 | -7.00 | **< 0.0001** | 0.31 |
| Ferritin | 325.0 [125.0 - 828.0] | 0.58 | -2.50 | 482.0 [245.5 - 993.5] | 0.55 | -2.71 | 0.181 | 0.61 |
| Globulin | 38.0 [34.0 - 42.0] | 0.07 | -8.19 | 40.0 [36.0 - 45.75] | 0.06 | -8.51 | **0.0161** | 0.57 |
| Glucose (Random) | 6.1 [5.4 - 7.4] | 0.57 | -10.61 | 6.95 [5.6 - 8.62] | 0.54 | -11.62 | 0.492 | 0.58 |
| Haemoglobin (Hb) | 135.0 [122.0 - 149.0] | 0.00 | -5.58 | 135.0 [122.0 - 149.0] | 0.00 | -6.06 | **0.036** | 0.44 |
| High Sensitivity Troponin I | 7.0 [3.0 - 23.0] | 0.25 | -3.64 | 37.5 [15.75 - 117.0] | 0.22 | -4.22 | 0.675 | 0.76 |
| Lactate dehydrogenase | 346.0 [270.0 - 537.0] | 0.53 | -3.86 | 383.5 [290.5 - 501.25] | 0.54 | -3.78 | 0.662 | 0.53 |
| Lymphocyte count | 1.0 [0.7 - 1.4] | 0.00 | -14.26 | 0.9 [0.6 - 1.22] | 0.01 | -13.59 | 0.961 | 0.45 |
| Mean cell haemoglobin (MCH) | 30.3 [28.8 - 31.4] | 0.00 | -8.70 | 30.5 [28.3 - 31.9] | 0.00 | -8.82 | 0.366 | 0.53 |
| Monocyte count | 0.6 [0.4 - 0.9] | 0.00 | -15.89 | 0.7 [0.4 - 0.9] | 0.01 | -14.66 | 0.127 | 0.54 |
| Neutrophil count | 5.4 [3.7 - 7.98] | 0.00 | -11.26 | 7.3 [4.57 - 9.8] | 0.01 | -11.79 | **0.017** | 0.61 |
| Neutrophils:Lymphocytes | 5.37 [3.2 - 9.82] | 0.00 | -11.10 | 8.11 [4.34 - 14.67] | 0.01 | -9.94 | **0.0006** | 0.62 |
| Platelet (PLT) count | 234.0 [183.75 - 294.5] | 0.00 | -4.13 | 216.0 [160.0 - 285.0] | 0.00 | -4.81 | 0.210 | 0.45 |
| Potassium | 3.9 [3.6 - 4.3] | 0.01 | -12.81 | 4.1 [3.73 - 4.6] | 0.03 | -12.97 | **0.0006** | 0.61 |
| Procalcitonin | 0.14 [0.06 - 0.38] | 0.41 | -9.22 | 0.31 [0.09 - 0.86] | 0.39 | -9.59 | 0.345 | 0.63 |
| Protein | 71.0 [67.0 - 76.0] | 0.07 | -6.87 | 70.0 [65.0 - 74.0] | 0.06 | -7.34 | **0.04** | 0.42 |
| Prothrombin time (PT) | 13.2 [12.4 - 14.48] | 0.14 | -10.17 | 13.95 [12.7 - 15.78] | 0.10 | -9.51 | 0.112 | 0.58 |
| Sodium | 137.0 [134.0 - 139.0] | 0.00 | -5.63 | 138.0 [134.0 - 141.0] | 0.00 | -5.79 | **< 0.0001** | 0.59 |
| Urea | 5.7 [4.0 - 8.52] | 0.00 | -10.67 | 10.2 [7.2 - 16.9] | 0.00 | -10.46 | **< 0.0001** | 0.77 |
| White blood cell (WBC) count | 7.35 [5.3 - 9.93] | 0.00 | -10.84 | 9.0 [6.4 - 12.3] | 0.00 | -11.39 | **0.016** | 0.61 |

### **Supplementary S2: Clinical Laboratory data on admission for evaluation cohort for the composite endpoint**

|  | **No admission to intensive care or death within 28-days** | | | **Admission to intensive care or death within 28-days** | | |  |  |
| --- | --- | --- | --- | --- | --- | --- | --- | --- |
|  | Median [IQR] | Missing (%) | Normalised Log Variance | Median [IQR] | Missing (%) | Normalised Log Variance | p-value | Common language effect size |
| APTT | 31.5 [29.2 - 34.3] | 0.16 | -8.44 | 31.0 [28.75 - 34.1] | 0.07 | -8.45 | 0.297 | 0.47 |
| Alanine transaminase | 24.5 [16.25 - 44.75] | 0.04 | -6.87 | 24.0 [15.0 - 37.25] | 0.01 | -7.44 | 0.19 | 0.48 |
| Albumin | 33.0 [30.0 - 37.0] | 0.04 | -8.19 | 29.0 [26.0 - 33.0] | 0.01 | -8.68 | **< 0.0001** | 0.30 |
| Alkaline phosphatase | 81.0 [66.0 - 114.25] | 0.04 | -6.24 | 93.0 [63.5 - 129.25] | 0.01 | -5.34 | **0.034** | 0.54 |
| Basophil count | 0.0 [0.0 - 0.0] | 0.00 | -22.66 | 0.0 [0.0 - 0.0] | 0.01 | -22.49 | 0.428 | 0.49 |
| Bilirubin | 10.0 [7.0 - 15.0] | 0.05 | -10.48 | 11.0 [7.0 - 15.0] | 0.01 | -10.44 | 0.165 | 0.53 |
| C-reactive protein (CRP) | 59.5 [17.75 - 121.25] | 0.00 | -5.53 | 102.0 [59.5 - 175.5] | 0.02 | -5.23 | **< 0.0001** | 0.66 |
| Clauss fibrinogen level | 5.1 [3.9 - 6.5] | 0.16 | -11.82 | 5.3 [4.0 - 7.25] | 0.07 | -11.83 | 0.147 | 0.55 |
| Creatinine | 82.0 [65.0 - 108.0] | 0.00 | -4.17 | 100.5 [74.5 - 175.0] | 0.00 | -5.27 | **0.008** | 0.64 |
| D-dimer | 919.5 [583.0 - 1776.0] | 0.30 | -2.84 | 1271.0 [850.5 - 3486.0] | 0.26 | -3.04 | **0.024** | 0.63 |
| Eosinophil count | 0.0 [0.0 - 0.1] | 0.00 | -18.61 | 0.0 [0.0 - 0.1] | 0.01 | -20.60 | **0.047** | 0.43 |
| Estimated GFR | 74.0 [55.0 - 89.0] | 0.01 | -6.39 | 56.0 [30.0 - 82.0] | 0.00 | -6.72 | **< 0.0001** | 0.37 |
| Ferritin | 293.0 [112.0 - 566.0] | 0.63 | -2.53 | 487.0 [228.5 - 1062.0] | 0.47 | -2.60 | 0.159 | 0.64 |
| Globulin | 38.0 [34.0 - 42.0] | 0.08 | -8.15 | 40.0 [35.5 - 45.0] | 0.05 | -8.44 | **0.024** | 0.57 |
| Glucose (Random) | 5.9 [5.3 - 7.25] | 0.62 | -10.55 | 6.9 [5.65 - 8.9] | 0.47 | -11.24 | 0.393 | 0.63 |
| Haemoglobin (Hb) | 134.0 [121.0 - 149.0] | 0.00 | -5.47 | 135.0 [117.25 - 147.0] | 0.00 | -6.05 | 0.216 | 0.47 |
| High Sensitivity Troponin I | 8.0 [3.0 - 22.75] | 0.28 | -4.21 | 22.0 [9.5 - 93.0] | 0.18 | -3.48 | 0.611 | 0.68 |
| Lactate dehydrogenase | 312.0 [259.0 - 410.0] | 0.60 | -3.83 | 419.0 [301.5 - 573.0] | 0.44 | -3.82 | **0.017** | 0.67 |
| Lymphocyte count | 1.0 [0.7 - 1.4] | 0.00 | -14.12 | 0.9 [0.6 - 1.3] | 0.01 | -13.89 | 0.938 | 0.45 |
| Mean cell haemoglobin (MCH) | 30.2 [28.8 - 31.4] | 0.00 | -8.60 | 30.6 [28.58 - 31.7] | 0.00 | -8.92 | 0.219 | 0.54 |
| Monocyte count | 0.6 [0.5 - 0.9] | 0.00 | -15.98 | 0.6 [0.4 - 0.9] | 0.01 | -14.91 | **0.105** | 0.50 |
| Neutrophil count | 5.4 [3.6 - 7.7] | 0.00 | -11.14 | 7.0 [4.4 - 9.8] | 0.01 | -11.71 | **0.0009** | 0.61 |
| Neutrophils:Lymphocytes | 5.1 [3.08 - 9.69] | 0.00 | -11.10 | 7.87 [4.11 - 14.5] | 0.01 | -10.23 | **< 0.0001** | 0.62 |
| Platelet (PLT) count | 232.0 [181.0 - 297.0] | 0.00 | -4.10 | 224.5 [171.5 - 285.75] | 0.00 | -4.48 | 0.518 | 0.48 |
| Potassium | 3.9 [3.6 - 4.3] | 0.02 | -12.73 | 4.0 [3.7 - 4.55] | 0.02 | -13.02 | **0.034** | 0.57 |
| Procalcitonin | 0.13 [0.06 - 0.31] | 0.43 | -17.06 | 0.31 [0.1 - 0.88] | 0.37 | -8.62 | **0.0028** | 0.66 |
| Protein | 71.5 [67.75 - 76.0] | 0.08 | -6.81 | 70.0 [65.0 - 74.0] | 0.05 | -7.24 | **0.009** | 0.42 |
| Prothrombin time (PT) | 13.2 [12.3 - 14.6] | 0.16 | -10.07 | 13.8 [12.7 - 15.2] | 0.07 | -9.79 | 0.575 | 0.57 |
| Sodium | 137.0 [134.0 - 139.0] | 0.00 | -5.56 | 137.0 [134.0 - 141.0] | 0.00 | -5.83 | 0.207 | 0.54 |
| Urea | 5.8 [4.2 - 8.55] | 0.00 | -10.58 | 9.0 [5.82 - 15.0] | 0.00 | -10.57 | **< 0.0001** | 0.68 |
| White blood cell (WBC) count | 7.2 [5.25 - 9.9] | 0.00 | -10.71 | 9.0 [6.02 - 12.2] | 0.00 | -11.33 | **0.0008** | 0.61 |

### **Supplementary S3: Laboratory test results according to survival outcome and grouped by age**

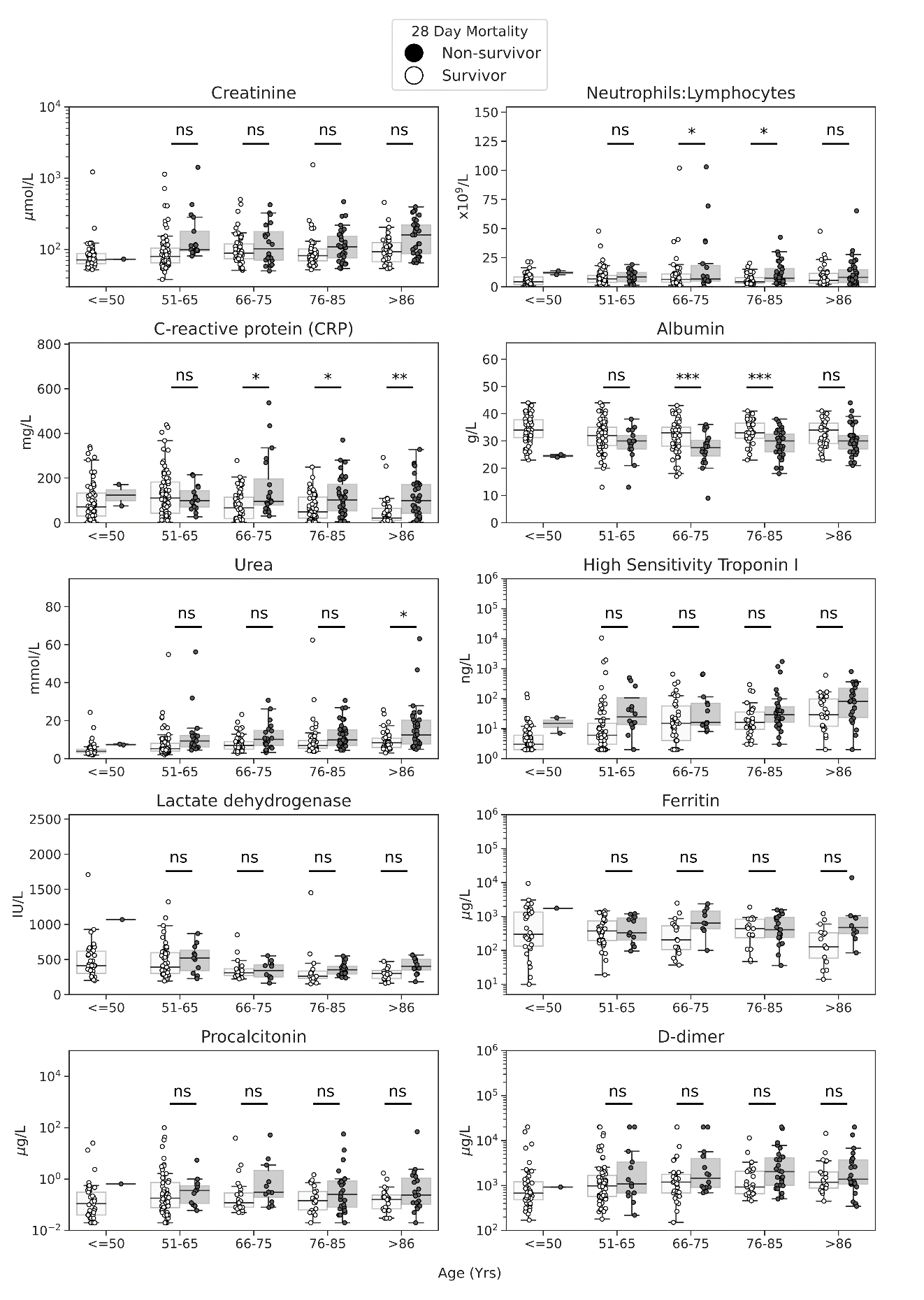
**Caption:** *Box and swarm plots showing the initial laboratory test results from laboratory-confirmed COVID-19 patients, grouped by age and 28-day mortality. Example variables considered from the components of the core laboratory test panel. * indicate level of significance, assessed by Mann-Whitney U test with correction for multiple testing: * p<0.05, ** p<0.01, *** p<0.005. **** p <0.001.*

Supplementary S4: Laboratory test results according to 28-day composite outcome, by gender

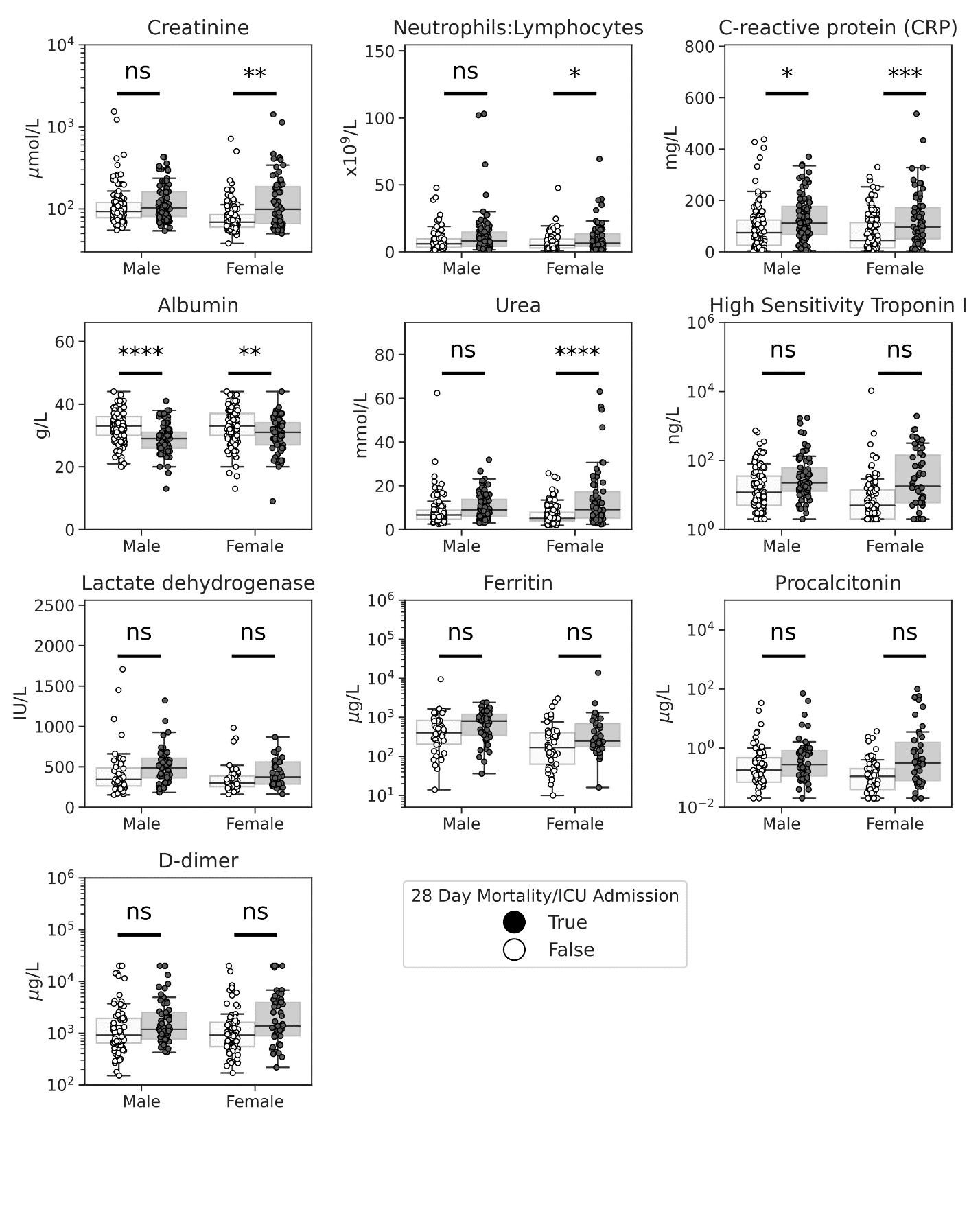

**Caption:** *Box and swarm plots showing the initial laboratory test result from laboratory-confirmed COVID-19 patients, grouped by gender and 28-day composite outcome (intensive care admission or inpatient mortality). Example variables considered from the components of the core laboratory test panel. Correction for multiple testing has been performed. * indicate level of significance, assessed by Mann-Whitney U test with correction for multiple testing: * p<0.05, ** p<0.01, *** p<0.005. **** p <0.001*

#### **Supplementary S5: Laboratory test results according to 28-day composite outcome, by age group**

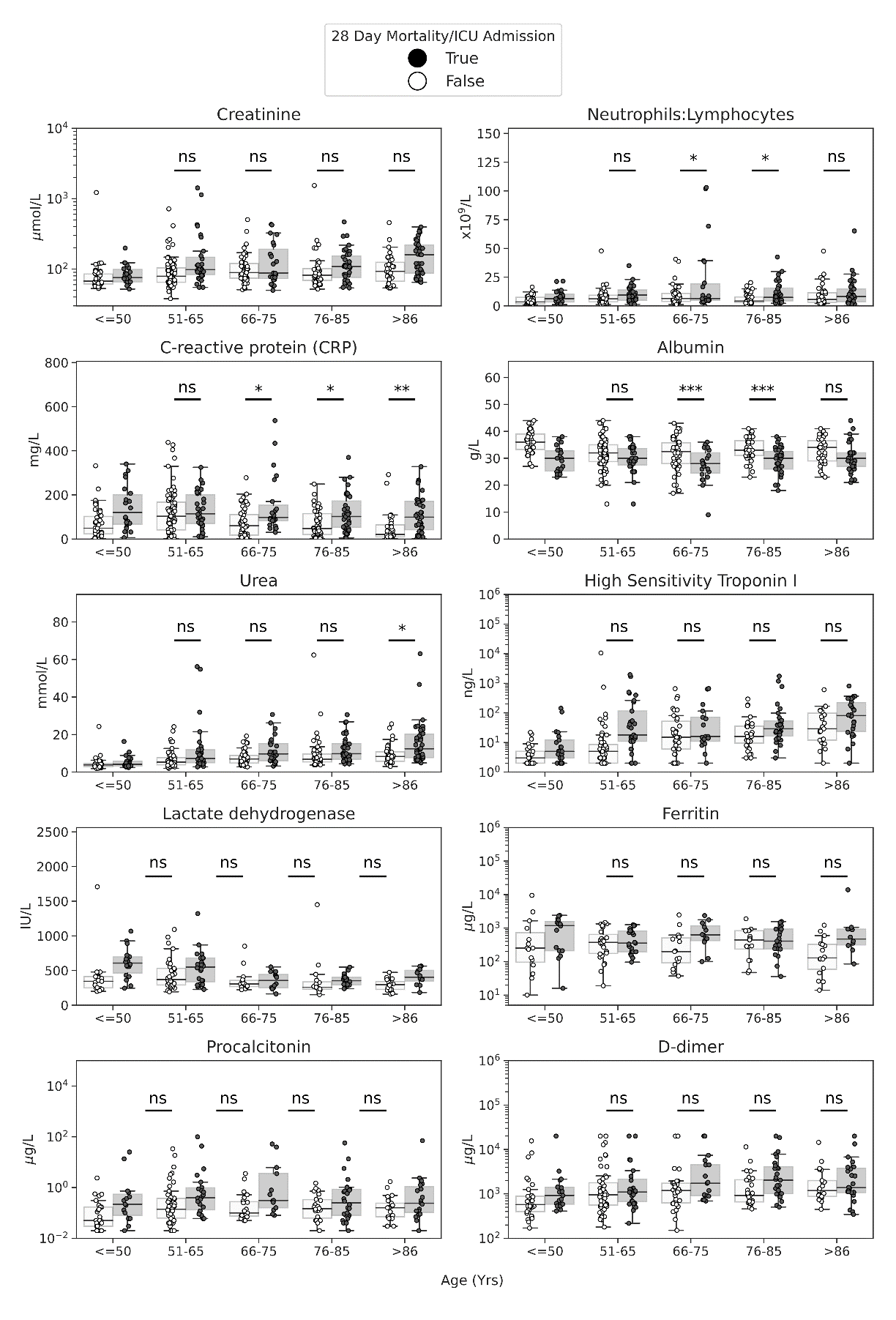

**Caption:** *Box and swarm plots showing the initial laboratory test result from laboratory-confirmed COVID-19 patients, grouped by age and 28-day composite outcome (intensive care admission or inpatient mortality). Selected variables considered from the components of the core laboratory test panel. Correction for multiple testing has been performed. * indicate level of significance, assessed by Mann-Whitney U test with correction for multiple testing: * p<0.05, ** p<0.01, *** p<0.005. **** p <0.001*

#### **Supplementary S6: Spearman-rank correlation matrix for assessing multi-collinearity**

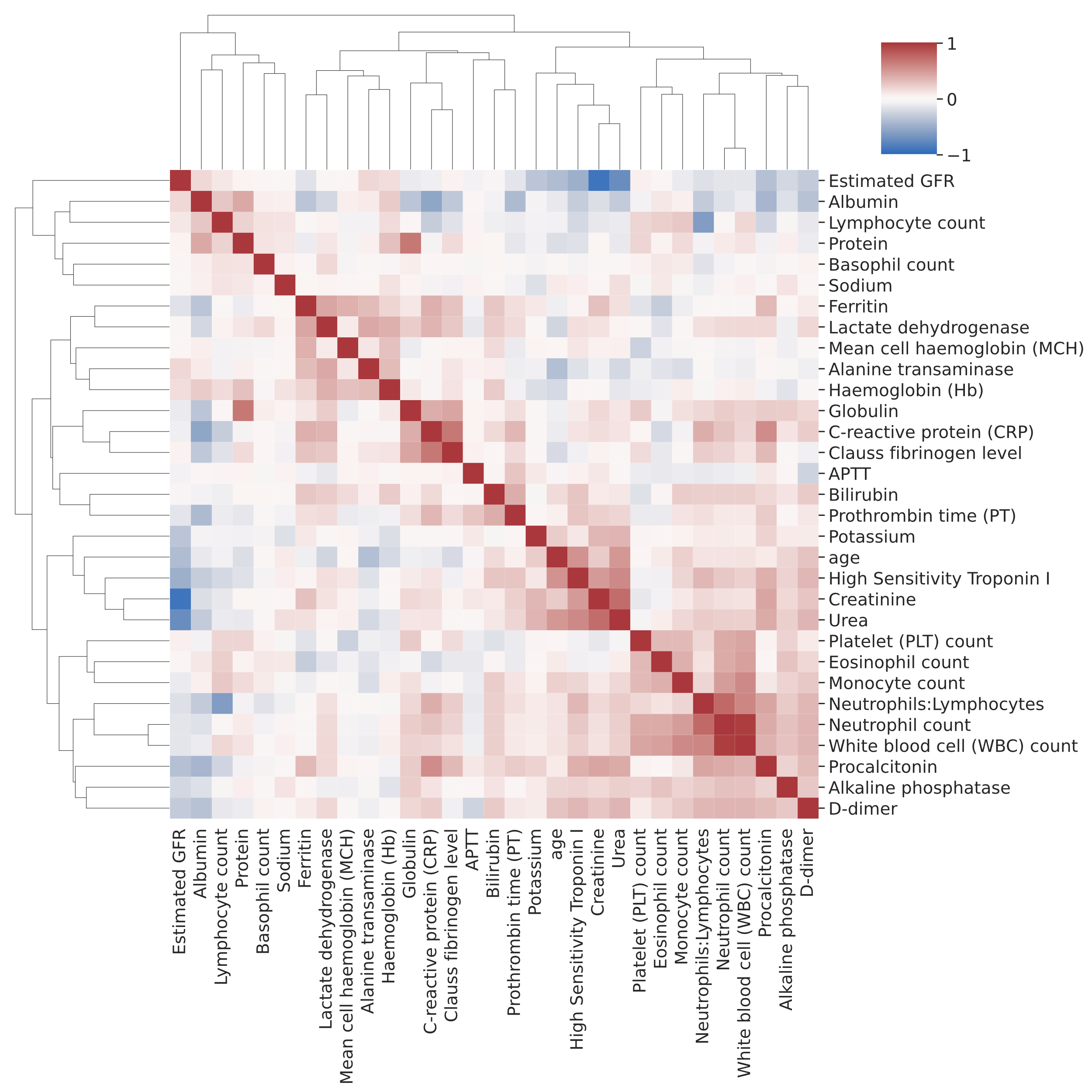

#### **Supplementary S7:** Area Under Curve analysis for prognostic performance using core and extended laboratory tests

| A: 28 day mortality | | |
| --- | --- | --- |
|  | **AUC** | **p-value** |
| Core features | 0.79 [0.67 – 0.91] |  |
| + D-dimer | 0.82 [0.71 – 0.85] | 0.15 [0.06 – 0.37] |
| + Ferritin | 0.79 [0.66 – 0.92] | 0.71 [0.49 – 0.94] |
| + High Sensitivity Troponin I | 0.78 [0.67 – 0.90] | 0.62 [0.49 – 0.75] |
| + Lactate dehydrogenase | 0.79 [0.68 – 0.91] | 0.23 [0.12 – 0.35] |
| + Procalcitonin | 0.78 [0.66 – 0.90] | 0.66 [0.35 – 0.96] |
| Extended panel | 0.78 [0.72 – 0.85] |  |
| B: 28 day mortality/ICU Admission (composite outcome) | | |
|  | **AUC** | **p-value** |
| Core features | 0.70 [0.56 – 0.84] |  |
| + D-dimer | 0.70 [0.56 – 0.84] | 0.61 [0.33 – 0.89] |
| + Ferritin | 0.69 [0.55 – 0.83] | 0.76 [0.47 – 1.00] |
| + High Sensitivity Troponin I | 0.72 [0.60 – 0.83] | 0.39 [0.23 – 0.56] |
| + Lactate dehydrogenase | 0.69 [0.56 – 0.82] | 0.31 [0.01 – 0.62] |
| + Procalcitonin | 0.70 [0.54 – 0.85] | 0.58 [0.36 – 0.81] |
| Extended panel | 0.69 [0.57 – 0.80] |  |

*(****A****) The performance of core features with sequential inclusion of additional extended biomarkers with 28 day mortality and (****B****) 28 day mortality or intensive care admission (composite outcome). AUC scores are reported as the average over five folds of stratified cross-validation with 95% confidence intervals. The reported p-values represent the significance of the additional component, averaged over five folds of stratified cross-validation with 95% confidence intervals.*

#### **Supplementary S8: Balanced accuracy (A), weighted F1 score (B), AUC score (C) and ROC curves (D) for models with sequential inclusion of extended biomarkers where the endpoint for prediction is the composite outcome of 28 day mortality or ICU admission**

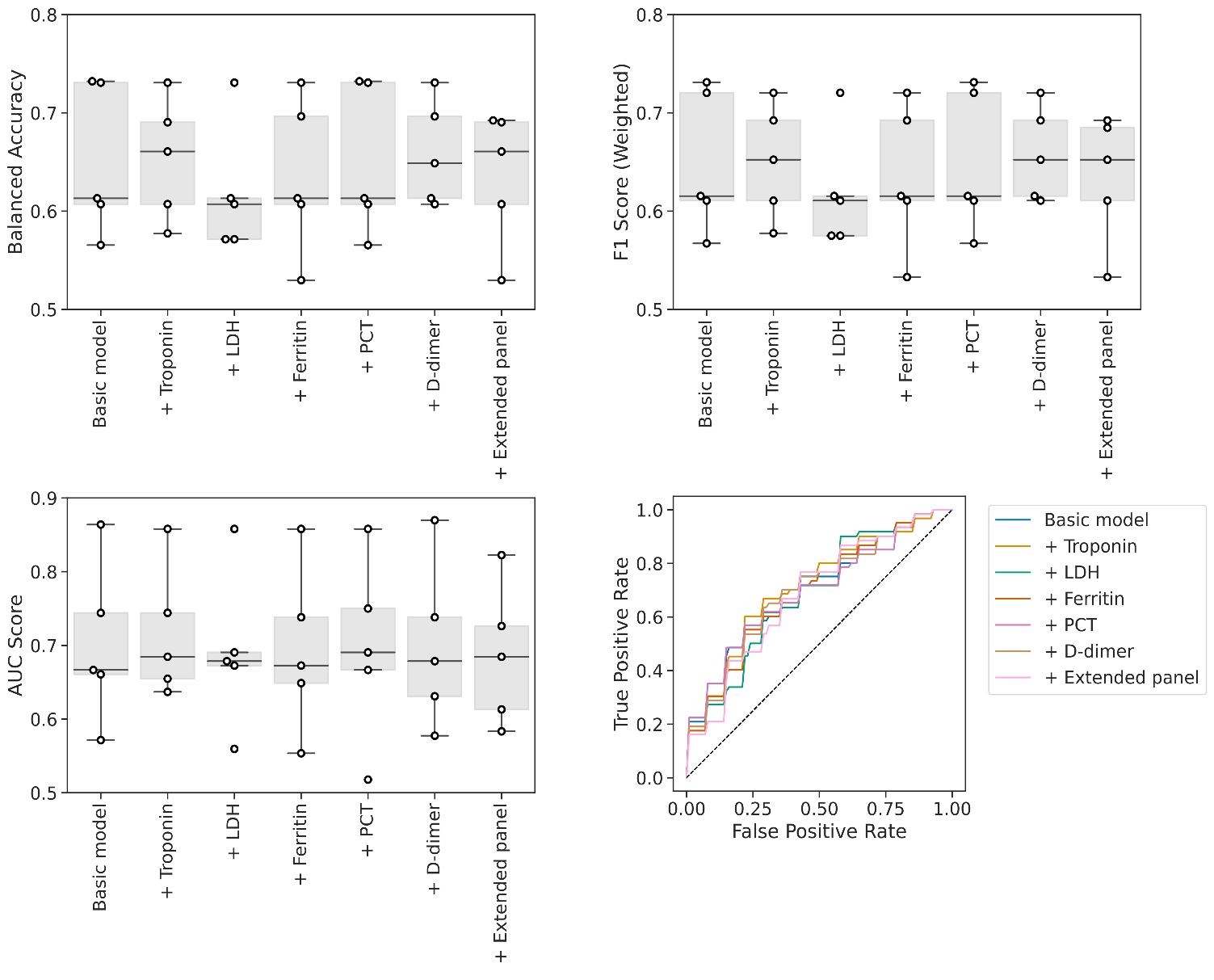

A

C

D

B

#### **Supplementary S9: Learning curves when predicting 28 day mortality, comparing training performance and cross-validated testing performance**

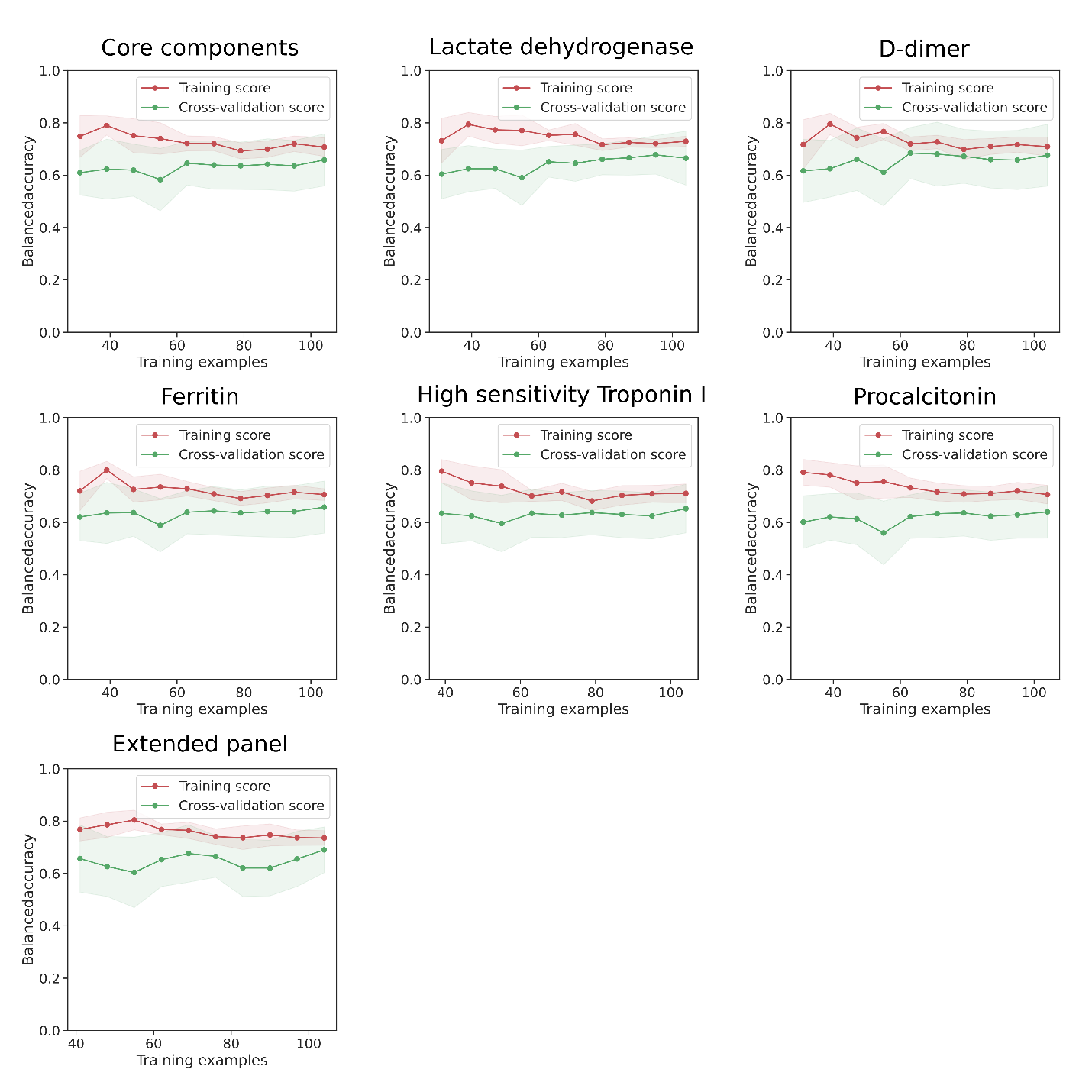

#### **Supplementary S10: Learning curves when predicting composite outcome of 28 day mortality or ICU admission, comparing training performance and cross-validated testing performance**

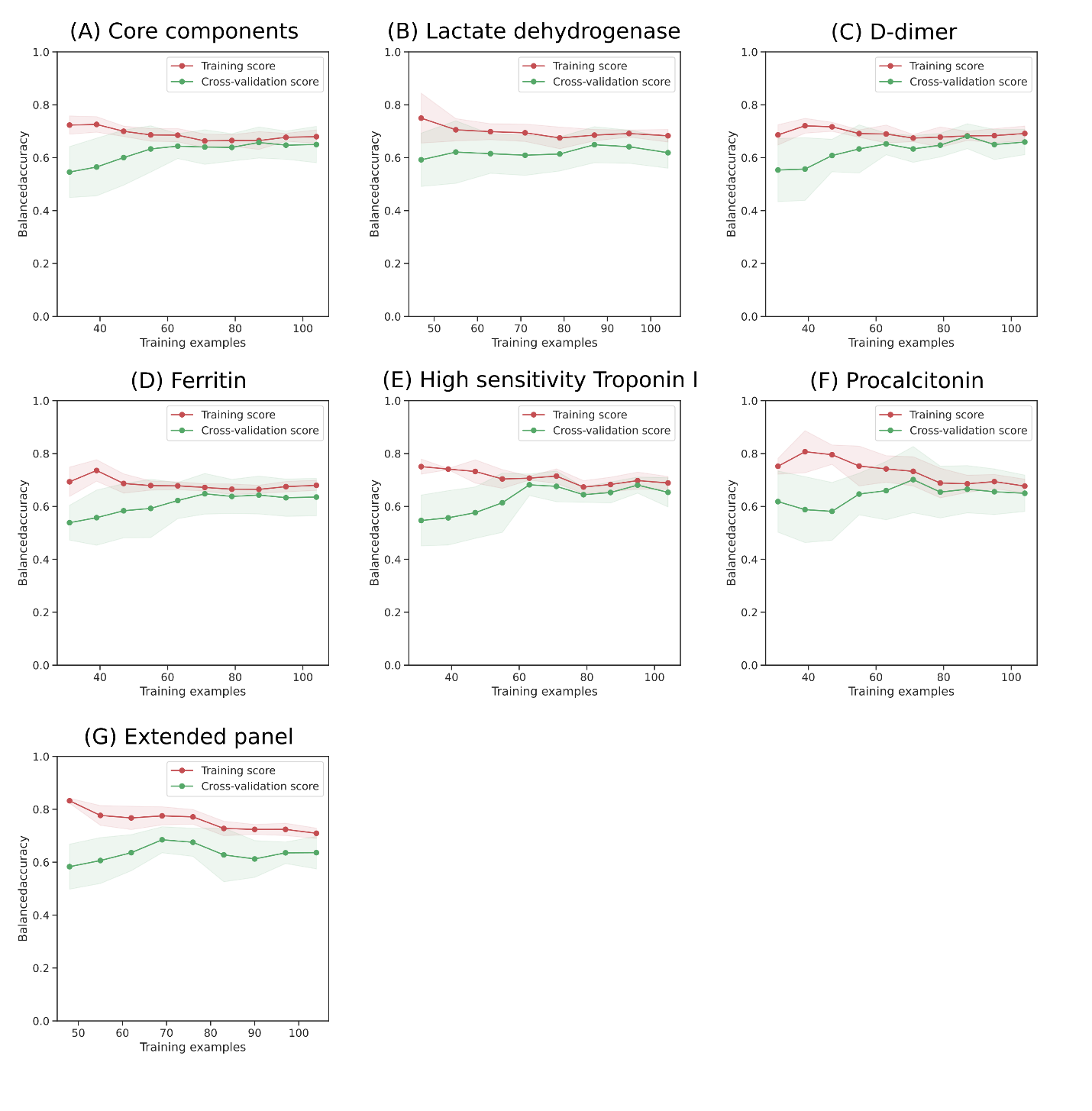

Supplementary S11: Threshold-performance curves for models with sequential inclusion of extended biomarkers where the endpoint for prediction is 28 day mortality.

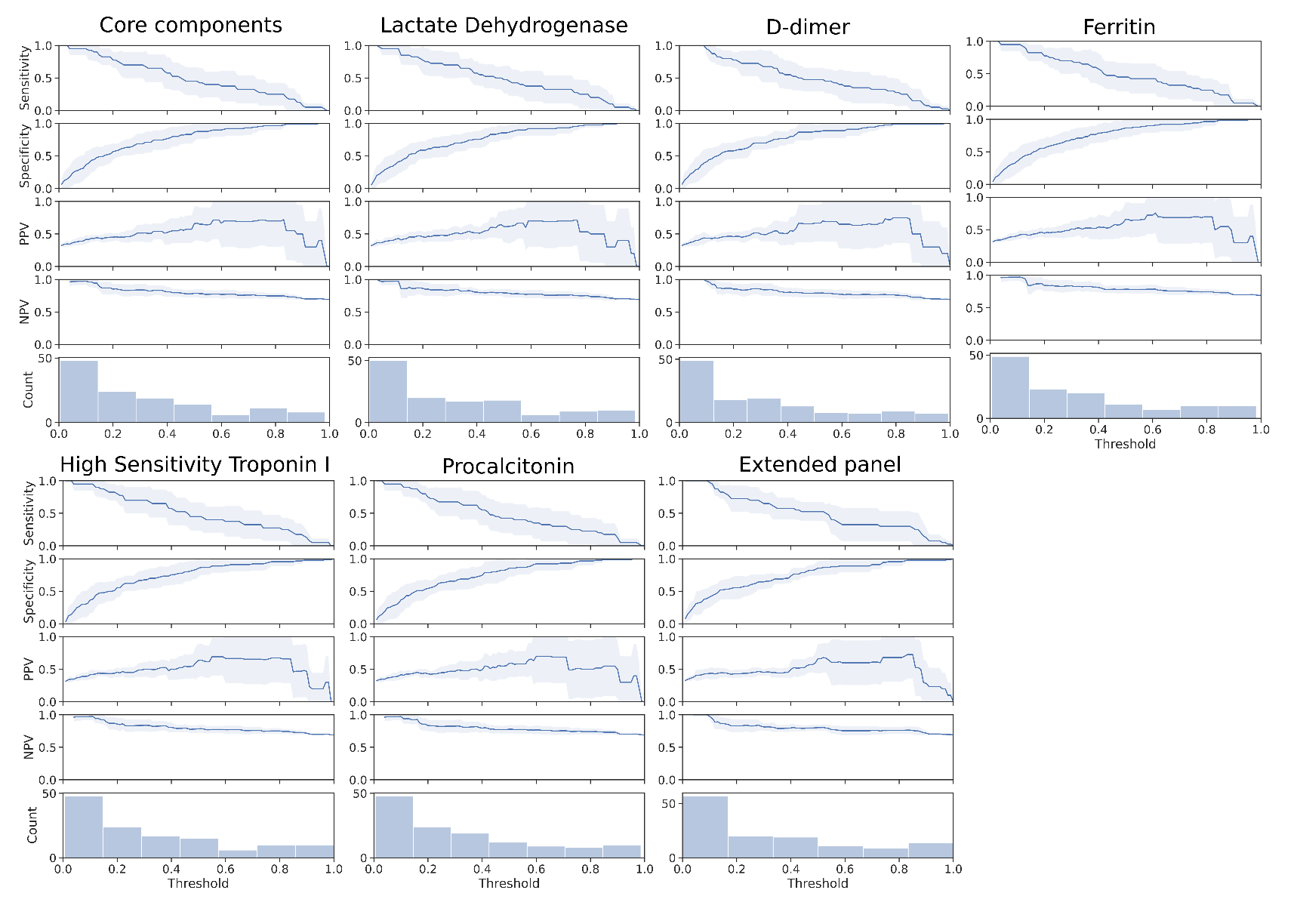
Plots show the quantity of predictions (across five folds of stratified cross-validation) over the range of probabilities of the positive case as a histogram. Above which sensitivity, specificity, positive predictive value (PPV), and negative predictive value (NPV) are plotted against the threshold of positivity.

Supplementary S12: Threshold-performance curves for models with sequential inclusion of extended biomarkers where the endpoint for prediction is the composite outcome of 28 day mortality or ICU admission.

Plots show the quantity of predictions (across five folds of stratified cross-validation) over the range of probabilities of the positive case as a histogram. Above which sensitivity, specificity, positive predictive value (PPV), and negative predictive value (NPV) are plotted against the threshold of positivity.

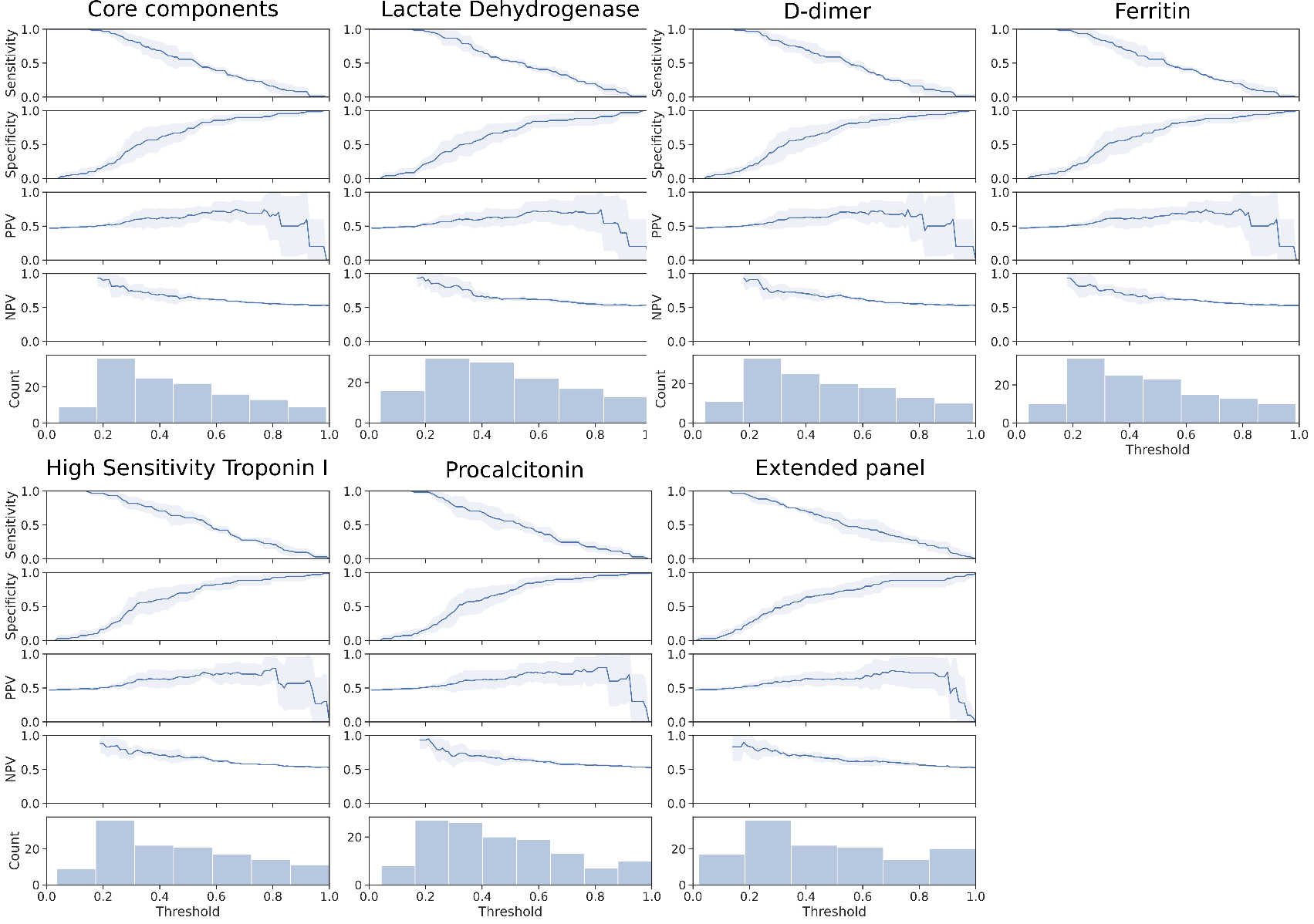

### **Supplementary S13: Local costs associated with laboratory testing panels**

|  | **Core** | **Extended** | **Estimated cost for NHS Wales requestor** |
| --- | --- | --- | --- |
| Full Blood Count | X | X | £4.31 |
| High sensitivity D-dimer |  | X | £16.40 |
| Ferritin |  | X | £4.80 |
| Urea & Electrolytes | X | X | £4.00 |
| Liver function tests | X | X | £4.86 |
| C-Reactive Protein | X | X | £3.27 |
| Lactate Dehydrogenase |  | X | £1.38 |
| High sensitivity Troponin I |  | X | £6.40 |
| Procalcitonin |  | X | £25.50 |
| **Total panel cost** | **£16.44** | **£70.92** |  |
